# Evaluation of the Family Liaison Officer (FLO) role during the COVID-19 Pandemic

**DOI:** 10.1101/2021.05.18.21256801

**Authors:** Luke Hughes, Lisa Anderton, Rachel M Taylor

**Author notes:** Correspondence: Dr Rachel Taylor.

## Abstract

**Objectives:** During the first wave of COVID-19 heavy restrictions were placed on hospital visitations in the United Kingdom. To support communication between families and patients a central London hospital introduced the role of the Family Liaison Officer. Communication within healthcare settings is often the subject of contention, particularly for patient’s families. During periods of crisis communication can become strained for patients and their families. We aimed to evaluate the rapid implementation of this role to provide guidance if it was required in the future and to explore the potential for this to become a standard role.

**Design:** Service evaluation

**Setting:** Single National Health Service hospital in London.

**Methods:** Semi-structured video interviews with a convenience sample of 12 participants. Data were analysed using Framework Analysis.

**Participants:** Family Liaison Officers (n=5) and colleagues who experienced working alongside them (n=7).

**Results:** Key themes were identified from the interviews pertaining to the role, the team, the impact and the future. Two versions of the role emerged though the process based on the Family Liaison Officer’s previous background: Clinical Family Liaison Officers (primarily nurses) and Pastoral Family Liaison Officers (primarily play specialists). Both the Family Liaison Officers and their colleagues agreed that the role had a very positive impact on the wards during this time. Negative aspects of the role, such as a lack of induction, boundaries or clear structure were also discussed.

**Conclusion:** The Family Liaison Officer was a key role during the pandemic in facilitating communication between patient, clinical team and family. The challenges associated with the role reflect the speed in which it was implemented but it was evident to those in the role and clinicians who the role was supporting that it has potential to help improve hospital communication, and the work of healthcare staff outside of a pandemic.

**Strengths and limitations of this study:**

- This was an in-depth evaluation of the Family Liaison Officer role from the perspective of those in the role and the clinical team who they were providing support.
- The sample included representation of the different disciplines who worked in the FLO role.
- The evaluation only represents the professional perspective and not the experience of the family.

## Introduction

COVID-19, a newly recognised and highly infectious disease and variant of the coronavirus family of viruses (which includes Severe Acute Respiratory Syndrome) was declared as a global pandemic in March 2020 by the World Health Organisation (1). In preparation for the first expected peak of the pandemic the National Health Service (NHS) mobilised emergency protocols to engage the surge in admissions. This resulted in many staff being redeployed to combat the developing pandemic, and restrictions being placed on movement throughout hospitals. Among these restrictions was a strict limitation for visiting patients admitted to hospital wards. This limitation was put in place with the aim to reduce the spread of the virus to both Health Care Workers (HCWs), inpatients and the general public, as well as protecting the available stock of Personal Protective Equipment (PPE) and COVID-19 diagnostic tests (2). Hospitals have a duty of care for their staff, patients and visitors; meaning that pragmatic decisions were needed to ensure policies of social distancing and control of the virus needed to be made. Given that the transmission of COVID-19 can transpire among asymptomatic patients, relying on screening tools to moderate visitation was generally disparaged (3). While it was recognised that limiting visitors would have an impact on the mental wellbeing of inpatients during this period, the global risk framework ultimately accepted the inherent uncontrollability of the situation as taking precedence (2, 4).

### Background

Despite the pragmatism of the visitation policies, restricting support from family or caregivers has serious implications not just for the mental wellbeing of patients but also their physical health (5). The incorporation of family members into a patients care has always been a complex issue in healthcare, as it requires a balance to be struck between the family’s need for information and access to their loved ones alongside the medical teams often time sensitive management of ill patients (6). Nevertheless, family members often play key roles in supporting patients in areas where healthcare staff do not have the time, such as at mealtimes (7).

Communication between the healthcare team and their patients’ families is a significant factor influencing patient experience, and one which is frequently the source of contention and complaint (8). Families frequently enter a space of liminality and powerlessness when a loved one is admitted to hospital and can feel further marginalised by the approaches taken by healthcare staff (9). However, in recent years a shift has been made towards family centred care, which focuses on communication and collaboration with the patient and family (10) and multidisciplinary approaches to patient recovery including the voice of the family for the development of short-term and long-term care plans (5).

Understanding family as a key element in patient experience and recovery in many ways conflicts with the limitation to visitor access resulting from COVID-19. These restrictions however are also vital for the protection of both HCWs and inpatients and for combating the spread of the virus in the community. In order to engage both these issues a new role was created during the initial peak of the pandemic. The Family Liaison Officer (FLO) was a role which was introduced at a central London hospital in the early stages of the first wave of the pandemic.

The aim of this evaluation was to assess the FLO role in order to better understand its efficacy and future. As this was a new role established in a short time frame during the early months of the pandemic it is important to understand what worked well, what did not work well and how it was experienced by both the individuals performing the role and those who worked alongside it. Recommendations on whether this role has continued value, both within and beyond the parameters of the pandemic, will be discussed, alongside improvements and recommendations for the purpose of the role formed through conversations with the FLOs and their colleagues.

## Methods

### Design

This was service evaluation using qualitative methods of data collection.

### Participants

The evaluation aimed to recruit all staff members who had been redeployed to the FLO role during wave 1 of the pandemic (n=9) and ten healthcare workers (HCWs) who had worked with them in the clinical area. The contact details of the FLOs were given to the researcher who invited them to participate through a group email. HCWs who had worked with the FLOs were likewise recruited via email, identified by the FLOs, and by the head of patient experience. The evaluation was conducted in accordance with Health Research Authority (HRA) guidance (11). The purpose of the evaluation was explained to the participants through a video call, who then were given the opportunity to ask questions. If they were happy to continue, they were asked to give a recorded consent. All participants were able to stop the interview at any time and were assured of anonymity and confidentiality.

### Data collection

Semi-structured interviews were held via virtual video sharing platforms. The interview guide was developed by the evaluation team through discussion with the patient experience team who managed the FLO role during its inception. There were versions of the interview schedule created in order to interview the FLOs and their colleagues. Interviews lasted approximately 40 minutes, but varied on participant.

Interviews were conducted by a member of the evaluation team with experience of qualitative interviews with HCWs and a background in mental health, in order to ensure participant wellbeing was a priority during the interview. Consent for the interviews to be recorded, for the use of quotations and the use of an AI transcription service were all provided at the beginning of the interview. Both the FLO interview and their colleague’s interview encompassed seven key topics: 1) the day-to-day running of the role; 2) positive elements of the role; 3) negative elements of the role; 4) effects on wellbeing; 5) efficacy of technology; 6) the relevance and future of the role; and 7) reflections on the role.

### Analysis

All interviews were transcribed using secure online AI transcription software. Once completed the transcript was anonymised, removing any identifiable information (names, job titles and ward locations). Participants were given option to review their transcripts if they desired. Data were analysed using Framework Analysis (12). After becoming familiar with the transcripts, a framework was developed based on the interview schedule but as interview text were indexed and charted into the framework matrix, this was developed to include additional themes as they emerged in subsequent interviews. The FLO and colleague interviews were analysed separately using the same framework and their perspectives were synthesised at the point of interpretation. This enabled divergences in their perspectives to emerge. The framework was created by two researchers, with one performing the analysis of the matrix while another independently reviewed the outputs for consistency.

## Results

Five FLOs participated in an interview. Three had a professional qualification in nursing and two came from a play specialist/youth worker background. Seven colleagues took part, consisting of two medical consultants on COVID-19 wards, three ward sisters and two volunteer service/patient experience team. Four key themes emerged from the interviews of both the FLOs and their colleagues: 1) The Role, 2) The Team, 3) The Impact and 4) The Future.

### The Role

The role theme had five subthemes: role activities; professional background; personal skills; role boundaries; and training and preparation.

#### Role activities

Two distinct versions of the FLO emerged through the interview process, hinging on the individual’s previous background. For the purpose of identification we will refer to these groups as the Clinical-FLO and Pastoral-FLO. Clinical-FLOs were redeployed from healthcare roles (i.e., clinical nurse specialists and research nurses) and put emphasis on liaising with patient’s families, providing medical updates and acting as a conduit between the medical team and the family unit. Clinical-FLO tended to be based on the higher need wards such as the High Dependency or Intensive Care step down units. These FLOs took a medical team approach to the role, attending ward rounds and liaising with the ward team. They would help to support patients communicating with their families, be it helping set up video/telephone calls or more interactive support based on patient needs. Clinical-FLO also helped with supplying provisions and comforts to patients, e.g. specific food items, though some did report they were unsure if this was a part of their role. However, the role was performed differently on each ward so there was a degree of variance in their approaches, with some more involved in the medical updating of families and dealing with the communication for end of life (EOL) patients or discharge, and others being more led by patient support.

> “I’m going on the ward round with them, knowing exactly what was going on with the patient and then sort of dividing up the work for who needs to be contacted by a doctor and who need who could be contacted by the family liaison officer. Whereas I know some of the other FLOs did it differently on different wards” (Clinical-FLO)

Another key element which some of the FLOs and their colleagues noted was the Clinical-FLOs having the knowledge and time to talk through patient care plans in more accessible ways than the ward team often had time for, ensuring patients were more involved and aware of their own treatment.

Pastoral-FLOs came from a non-clinical background (i.e. play specialists and youth workers) tended to focus more on holistic and pastoral patient care. They tended to liaise with families but did not provide any medical updates as they did not feel confident or best placed to do so. However, they did provide social updates in terms of patients’ comfort and how they were spending their time. The Pastoral-FLO put emphasis on keeping patients engaged, comfortable and stimulated during their admission. These FLOs spoke mostly of spending time with patients by the bedside to help mitigate loneliness and boredom and providing elements of pastoral care (i.e. decorating a patient’s room).

> “We would go in and get the newspapers and do newspaper round, then we do a little tea round and then sort of see who wanted to speak to their families and sort out the iPad, and then by that time it was, you know, lunchtime and then sitting with them, helping them if they needed help eating their lunch, because a lot of them did need help with that and then in the afternoon, we tried… to do at least sort of one like focused activity a day. So whether that was…bingo, we did a ward quiz…painting…and they were making stuff for their grandkids. And we tried to sort of do one activity a day, so they had something to focus on in the afternoon” (Pastoral-FLO)

In some ways the Clinical-FLO may have been more focused on the families and communication element of the role, while the other FLOs were more led by patient experience. Regardless of background all of the FLOs were shown to play key roles in building relationships with patients and families which were viewed as incredibly useful by the ward staff. Likewise FLOs of either background took on important roles in supporting communicating with families, including EOL patients, and advocating for the needs of their patients. The colleagues of the FLOs all found the role very useful and complementary to their ward teams. In particular, colleagues of the Clinical-FLO found that having a consistent touch point for family communication was time efficient for both the families and the ward team alike.

> “It was very good for time efficiency, to actually spend time phoning each family member… was really quite time consuming, big time efficiency saver for us as doctors. So on a practical level that was really important for us” (Colleague)

Similarly having a member of staff with insight into the patient’s individual home lives was useful in terms of care planning and discharge. This was reported similarly by the colleagues of the Pastoral-FLO. However, while these FLOs did not handle the medical updates their colleagues found the social updates for families very helpful in easing anxieties, and that engagement of patients throughout the day had a positive effect on patient morale and subsequently the morale of the ward staff.

> “Having that insight from the FLOs was really valuable. They would pick up on all sorts of things that, you know, we just wouldn’t either get from the patients because they don’t feel it’s relevant to tell the medical team about a certain kind of social situation or something… we found out a lot more about their home situations” (Colleague)

#### Professional Background

For the Clinical-FLO their backgrounds were viewed as essential for understanding patient care, helping patients to understand their own treatment plans and being able to give meaningful updates to the family. They felt not having this background would make this role difficult. The FLOs who were originally senior nurses felt that even nurses who did not have as much medical experience were likely to struggle. In particular, it seemed having a background in the ward speciality was key; a FLO placed in a surgical ward with a surgical background for example felt it would be difficult to work there without insight into the minutia of the ward’s functions.

> “I felt like if I hadn’t had those experiences, it would have been very difficult to explain to patients what was happening” (Clinical-FLO)

This was reflected in one FLO’s experience when their ward reverted from a COVID-19 ward to its original area. Without much experience in this area they struggled with how to support families with this unknown area of work.

> “It was quite difficult to have the conversations because I didn’t really know what the tests were, things I hadn’t heard of before” (Clinical-FLO)

It was also noted that having some level of seniority within the trust was useful. Key professional skills that Clinical-FLO felt were useful included risk assessment and accountability, awareness of how to communicate appropriately with families, as well as the ways to escalate concerns of either patients or families within the correct channels.

> “From my experience, and I think as a nurse, you know, everything you do, you’re accountable for. You would risk assess it. If someone asked for something and you think ‘Oh, actually, this wouldn’t be right to do this to this patient’… you’d have to know how to escalate them properly. I suppose as a nurse, you know all of these things” (Clinical-FLO)

On the other hand, the Pastoral-FLO did not feel confident with regards to giving medical updates to families. These FLOs brought their skills of engaging patients to the role, providing holistic and pastoral care, helping with meals, sitting with patients and giving them company and emotional support, and trying to keep them engaged by having daily activities to prevent boredom and loneliness. These FLOs seemed best-placed for care of the elderly as opposed to be being place on the more intense medical wards such as the High Dependency Units or surgical wards. Their background in play and youth engagement was shown to have a high level of transferability to working with patients experiencing dementia or delirium.

The colleagues of the Clinical-FLO agreed a medical background was an essential element of the role, and that a level of seniority within the trust was desirable. It was noted that similar roles had been unsuccessfully attempted before, using lower banding and inexperienced staff. Most agreed that while a nursing background was not always necessary; having some clinical knowledge would be important if the role was to be implemented as standard. This was felt as important in order that the FLO understood patient’s circumstances and health status, to give meaningful updates to families, help patients to better understand their treatment plans, and to have awareness of what level of detail was appropriate to communicate to the family. However, it would also be important for the FLO to have some experience of emotional support. Colleagues of the Pastoral-FLO felt that the role highlighted a missing element of geriatric care. They did not feel that giving medical updates to families was as essential as giving social updates for this client base who tend to be in hospital more long term, with less immediate risks.

#### Personal Skills

The important personal skills for a FLO were unanimously agreed upon by Clinical-FLO, Pastoral-FLO and their colleagues. It was agreed that being a good communicator was essential, particularly knowing when to speak and when to listen. Being a caring, compassionate, empathetic, flexible, and a dynamic person were also the main attributes which were noted to be important.

> “It’s really just the values isn’t it is the innate kindness, some patience, its understanding and kind of just being empathetic” (Colleague)

Some FLOs (both clinical and pastoral) as well as their colleagues, noted having skills in conflict resolution and de-escalation were very important as they were often dealing with very distressed family members, particularly with regards to adhering to visitor policies. Resilience was also noted as a key skill which was needed given the emotive nature of working with critically ill patients who were frequently distressed, isolated and anxious.

> “Good negotiating skills, because we do get relatives who are very angry about not being allowed in to visit. And we have had situations particularly out of hours, where it’s been really uncomfortable” (Colleague)

#### Role boundaries

FLOs appeared to struggle initially with the role, as there were no clear boundaries, duties, or line management for them on the wards. Many felt they had been given little preparation before being thrust into the role.

> “There wasn’t a lot of organisation, it was just sort of… thrown into it to see what we could do” (Clinical-FLO)

Given the novelty of the role, it was not clear to many staff what the FLO did, and without a clear definition, this also made it difficult for the FLOs themselves. Many of the wards were made of teams comprising of newly deployed staff who were unfamiliar with their co-workers or the clinical setting, and with no induction or introduction the FLOs found it difficult to establish themselves.

> “They don’t know actually what your role is. They think you’re looking after the patients. So instead of the nurses looking after the patient and us being present, we had to explain that over and over, I felt like every time I had to explain what my role was…what I was doing here” (Clinical-FLO)

Some noted that other staff would frequently ask them to carry out tasks which were not outside their professional remit but were outside the parameters of the FLO role (i.e. those with nursing backgrounds being asked to check control drugs).

> “I was asked to come and check control drugs. So then I had to say, ‘Well, look, I have been non clinical, and this is my background. And I don’t think I should be checking control drugs with you, even though if it is just a matter of just counting and signing and checking.’ So I think they found it a bit difficult to understand this” (Clinical-FLO)

Setting those boundaries seemed to be an individual task, meaning some FLOs carried out some activities that others did not, which could potentially have resulted in inconsistent care. Many described feeling distant from the rest of the ward team, and not knowing where exactly they fitted. The lack of a clear line of duty for the role also meant that some FLOs were unsure if their duty of care was primarily with the families or with the patients, and though they all did naturally begin to focus on both the patients and their families, it was unclear in the initial stages of their deployment.

> “I ended up supporting the patients to some extent as well, because I was working five days a week and I was seeing the patients frequently…I got to know the patients and the families quite well. And I don’t know that that was necessarily part of the role. It just sort of happened.” (Clinical-FLO)

This ambiguity could be stressful for the FLOs in a time that was already generally difficult for many staff. Some mentioned conflict with management who they felt were taking a hard line on visitation policies, and felt they clashed with management in order to advocate for their patients. A clear benefit for these FLOs was meeting with other FLOs to discuss their roles, which allowed them to grow their skills and methods as a group.

> “Especially at the beginning to see how each other were doing it because none of us really knew what we were doing. And there was no job, no real job plan.” (Clinical-FLO)

Colleagues of the FLOs noted boundaries were not always clear for them either. In particular they did not know to what extent a FLO should be expected to communicate medical information to families, such as breaking bad news. The majority of ward-based colleagues felt this was something that was the responsibility of the doctors. Overall, most medical staff believed that the more serious medical updates should come from the medical team and not the FLO, but that a certain level of routine medical updates on patient’s status were greatly appreciated by families and could be effectively provided by the FLO.

> “It is good for efficiency for the doctors, but at the same time, I’m not sure it can completely replace the primary medical team communicating with the family, especially around the more difficult and complex decisions” (Colleague)

Ethical boundaries were discussed by both FLOs and their colleagues. For example, there were no guidelines on how to react in some situations regarding privacy for EOL patients who were communicating with families online, where families were bringing other family members to join the call without the patients consent. Likewise, clarity was needed on what the safeguarding for both staff and patients would be in scenarios where families wanted to speak with a patient but a patient did not want to speak to the family. In order to protect the rights of all parties involved some guidelines on this would need to be introduced.

> “You put them on and people would be dying and then they’d be dialling in lots of other people to the call and sometimes I don’t know as a nurse that just felt a bit inappropriate because there was sort of no consent process there for this dying patient” (Clinical-FLO)

#### Training and preparation

No training or preparation was provided prior to being redeployed into the FLO role. Some felt that there would have been no time, while others wished they could have been prepared more. Some Clinical-FLO felt that in the future non-clinical staff could be given enough training to help them understand the basic clinical knowledge, but not to the degree that healthcare staff would have. Overall, redeployment was a stressful and chaotic time for everyone. Many FLOs felt very unprepared for their redeployment and were given little to no time to prepare for the role before it began.

> “It was very chaotic at the start, so it was very quick to happen. So it was sort of … I got a phone call, I think on the, it was like Monday or Tuesday and then the role started two days later. So it was very quick” (Clinical-FLO)

When the role began FLOs were given no clear direction. However, for those who remained within their original clinical team, establishing the FLO role was much easier.

> “There was no sort of induction or anything; it was just sort of an idea. So you’re going to be a family liaison, here’s a phone, now go in contact with patients’ families. And that was basically it. And there was no other instruction of what to do, really. And nobody on the ward had any idea of what we were meant to be doing.” (Clinical-FLO)

Many struggled with the not having a timeframe to know how long they would be redeployed for, and when the role ended many felt there was no clear transition back into old role which was equally as difficult. However some agreed given the circumstances there was not much that could have been done.

> “I think the uncertainty of being redeployed like that that was a really strange feeling and not knowing when I’d go back to my other job and how long redeployment would last so it’s quite, there was quite a lot of feelings” (Clinical-FLO)

Overall, they all felt they had positive and meaningful experiences working as FLOs. Their ward colleagues agreed that they had little preparation to receive the FLOs on the ward, without a specific brief or formal induction of the role. This caused some initial confusion, and it was felt that more structure for the role was needed for them to know how best to lean upon the FLOs, and utilize them to their capacity. For example one colleague noted that they were unsure how much they needed to communicate with the FLO throughout the day which lead to some miscommunication of tasks;

> “Sometimes it wasn’t entirely clear exactly when she was going to call which family in which order and obviously things are quite fluid and dynamic and medical wards and some patients are getting sick and it’s more of a priority to call that family first. So we maybe could have structured it a bit more to sit down with her and go through who we felt were the priorities. We perhaps left that to her” (Colleague)

Overall, all of interviewed colleagues felt the role had been a hugely positive success, but agreed that for the sake of both the ward team and the FLOs a formal induction to the role would be necessary next time. This would have diminished responsibility of the FLOs developing and establishing the role themselves, while simultaneously performing it and dealing with the difficulties inherent to redeployment.

> “When they arrived on the ward, we couldn’t really tell them exactly what we wanted because we weren’t sure ourselves. But they managed to work out and do some trial and error and work out the best way to do things” (Colleague)

### The Team

Clinical-FLO redeployed to wards that consisted mainly of redeployed staff tended to feel more isolated. High turnovers of staff, not having a defined role, no induction to the team and unclear boundaries in the FLO role initially made it difficult to bond with ward teams.

> “When you got to the ward it was a bit less just a bit less organised people weren’t expecting us…that was difficult because I think…you didn’t really have like a team to slot into…you felt like a bit on the outside looking in…But after a month or so I settled into it slightly…I wasn’t part of the nursing team or the medical team or the allied health professionals. So it was difficult to know where to sit” (Clinical-FLO)

Many of the Clinical-FLO found good support in the other FLOs who they met with on a semi-regular basis and had a WhatsApp group in order to stay in contact. The Pastoral-FLO, who remained part of their original team, found it easier to access team support.

The colleagues of the FLOs had empathy for them, understanding that new roles can be difficult to instil in a hospital as there is often aspects of territorialism imbedded in ward cultures, which are difficult to break. Without an introduction to the ward it was difficult for staff to know how to rely on the FLOs, and some staff felt the FLOs were taking on tasks they believed to be their own.

> “They were apprehensive at the beginning because they were coming in and saying let me do this for you. Let me do that for you. And you know, people can be a bit territorial” (Colleague)

While FLOs did become an integral element of the team, there was a degree of separation from their co-workers on the ward. FLOs who remained on their original wards with original team transitioned into the role much better and served to help support newly deployed staff to settle in. Many FLOs were seen to not only emotionally support patients and families, but to also take on supporting the staff on the wards. This may have been related to the types of personality that was generally placed in the FLO role.

> “They were quite junior nurses where I was working, and who were - not that they were unhappy, but more it’s like a fear, you’re coming to the unknown and having to do all these skills…So in terms of those as well, I had to support them.” (Clinical-FLO)

### The Impact

The FLO role had an impact on staff wellbeing, and patient and family experience.

#### Staff wellbeing

COVID was a stressful time for everyone, where a period of adrenaline proceeded widespread emotional exhaustion following the peak. Many FLOs struggled with the liminal space of their redeployment and not having any clear plan about when they would return to their old roles. Some FLOs felt isolated and would have liked more clinical supervision. The work with COVID-19 patients was noted to be particularly draining. The unknown nature of the virus and the critical level of sickness that affected some patients meant many patients experienced a turbulent medical journey. Given the nature of the FLO role meant that staff became quite close to patients and their families, they inevitably were taken along on this journey as well. FLOs from both clinical and non-clinical backgrounds were no strangers to complex medical journeys or EOL patients. However, the mercurial nature of COVID-19 saw patients vacillating between extremes of health, which had a notable impact on the staff.

> “When they were discharged, I felt really emotional, which I was quite confused about how that felt. I thought I should feel really happy. But I think it was just, you know, you’ve seen people really sick and then they bounce back and you do have that really close contact with the family. So you feel for them, you feel empathy” (Clinical-FLO)

Elements of burn out were noted in many of the FLOs, particularly with regards to the surge of patient numbers, creating a revolving door scenario of patients and families experiencing extremes of undulating health during their admission.

> “I think it got to around end of May, June, and I think I crashed – we said we’ve got to do it and then actually it all caught up and I was like, that was a lot” (Pastoral-FLO)

Some staff found support more useful in their own teams, while others did not feel comfortable in group psychology sessions with their team or attending sessions from home due to their family being present. The Clinical-FLO often found support in the other FLOs while the Pastoral-FLO were largely contained within their own team which had pre-existing support sessions; they frequently engaged in these on the ward. Fears over transmitting the virus to family members were also a cause for concern. Some noted that during the initial stages of the pandemic when it was ‘*all hands on deck’*, there was no room for seeking psychological support but when numbers of patients reduced considerably, staff felt stranded in a liminal space waiting to be deployed back to their original roles. This was the time when they experienced more complicated emotions.

> “The thing that caused me the most stress was not knowing how long I’d be redeployed for…and like knowing at any stage, because I remember being really anxious before I was redeployed, and then then I was…for about two weeks I felt ok…then I thought, oh, gosh, this is gonna happen again, because I need to get back to my old job…that’s more about redeployment than the FLO role to be honest” (Clinical-FLO)

Colleagues of the FLOs also agreed that the pandemic had been a tough and exhausting period, and that FLOs had the added emotional burden of dealing with distressed family members. They also felt most support had been internal within their teams. Some colleagues felt that the role of the FLO took on some of the extra burden of emotionally supporting patients which facilitated time for the ward team up to be more engaged in time sensitive care. Other colleagues maintained that even with the presence of the FLO it was inevitable to take on the emotional burdens of patients.

#### Patient and Family experiences

While patient care was more central for some FLOs than others, all FLOs discussed supporting their patients. The level of death was difficult for them to deal with, and many were emotionally involved with their patient’s. While visitors were not allowed in the hospital, changes were made to the rules for patients who were EOL. This was something many of the FLOs were instrumental in advocating as they were acutely aware of the patient and family’s enhanced psychological needs at the time.

> “I was trying to argue the case where, you know, there are there should be some exception to the rule because of patient psychology… where we could do it safely where the patient were on the site.” (Clinical-FLO)

As noted previously, patient engagement and entertainment was much more of a focus for the Pastoral-FLO; a colleague working in elderly medicine noted:

> “They really sort of developed this thing where they organised cognitive stimulating activities, and that were sort of age specific. So things like, and they had like tea parties, and like, bingo, and quizzes, and it was really imaginative. And I suppose it was applying what they normally do in paediatrics to geriatric care for the purposes really, and keeping people cognitively stimulated…patients were happier and staff were happier as a result” (Colleague)

Clinical-FLO also engaged in patient support and engagement but perhaps not to the same degree. All FLOs noted the difference even small comforts could bring for a patients experience of hospital care and felt the role generated many experiences of meaningful work. Colleagues of the FLOs found the role invaluable for patient-centric care. Having a role specifically mandated for giving patients the time and care that the ward staff simply did not have time for was seen as hugely beneficial for patients, family and staff. They found that the FLOs built up good rapport with patients and that they were an essential locus of insight into patient’s individual circumstances, nuances and needs.

In terms of family support, FLOs perceived that COVID-19 was an extremely distressing time for families of ill patients due to both the risk of death from the virus and the strict rebate on visitation.

> “Dealing with difficult…distressed patients dealing with distressed relatives, and you know, some relatives expectations are one call a day, one call a week, some relatives…we have them queuing outside the hospital waiting…their level of anxiety was, you know, off the scale “ (Colleague)

FLOs felt that anxiety was found to be particularly high if they were not stationed on the ward over the weekends, leaving families with extended periods without substantial updates. The medical updates were very important to some families more so than others, but all families appreciated being able to contact a FLO and knowing that person was there to reach out to if needed.

> “The FLOs had much greater insight into kind of family dynamics and what the patients were like outside of hospital and just they had a bit of time to kind of chat to the families and, and get to know the patients a bit better” (Colleague)

FLOs were instrumental in helping families communicating with patients throughout this period, which was particularly salient for EOL patients. Colleagues of the FLOs noted how the normal means of communication between hospital staff and families simply did not function in the current circumstances. Many pre-COVID methods of communication on hospital wards tended not to be family focused; FLOs reflected that staff often promised to return to speak to them but invariably became busy, so the patient and family were left with unresolved anxiety and stress.

> “One thing that we find with nursing and we often say…’I’ll get back to you in a few minutes’, but a few minutes is normally a few hours, because time just vanishes on the ward and so they then I think, from my perspective, families were given a really robust system of being contacted on a regular basis and given that that time schedule of when they’d be called or you know, and it worked with them and what their needs were and their anxieties” (Colleague)

FLOs were important for bridging this gap in care. The role gave families structure of when and how they would be contacted and a person they could contact when needed.

> “The biggest frustration of relatives certainly at the moment is the nurses change each day, because they’re all doing shifts, so they don’t get to speak to the same nurse twice, and the nursing charge phone, which is currently their only contact with the ward is used as kind of a clinical liaison phone, so has multiple people trying to get through at all times and relatives can’t get through whereas there was a dedicated phone that the FLOs had, the relatives knew…they would get to speak to someone who knew their patient “(Colleague)

Families differ in how much contact and updates they want, and the FLOs were very able to identify and cater to these needs. Likewise the Pastoral-FLO provided social updates to families, which were also useful at easing anxieties, helping families to see patients were comfortable and even in some cases enjoying their experiences on the ward.

> “The patients really enjoyed it and the staff, and really liked to be on the ward as well, because like, it was a sort of nicer environment” (Colleague)

### The Future

All of the staff redeployed to the FLO role felt it was an important role and should be continued where possible. Consistency of care was identified as a clear benefit, by having the same staff based on the same ward in order to help build rapport and trust with patients, family and clinical staff. Many experienced meaningful moments in their role and connections with families and patients. For example a Pastoral-FLO described the meaningful experience of an elderly patient who thrived in the care of the paediatric team.

> “We had a patient that was that they said with palliative…they took him off end of life once we started spending time with him daily, like we’d spent a good couple of hours, keeping him busy, like doing his hair in the morning, like getting him back into a routine. He left the ward six weeks, he said that they should rename the ward a holiday camp - he thought it was really great.”(Pastoral-FLO)

Colleagues of the FLOs felt the role was valuable. They agreed having a consistent member of staff was useful for families to contact and build a relationship with. Colleagues felt this could be a huge improvement to hospital communication, which was often an area of contention. The typical routines of ward staff within a hospital can conflict with the interest of families, such as repeatedly changing roster of nurses which can be frustrating for families. When FLOs returned to their original roles, ward staff were failing to keep up with the demand of the communication needs of families, which speaks for the efficacy of the role in of itself.

> “We’re finding increasingly that the communication with families is really difficult at the moment when visiting isn’t back up…I think actually the role itself was so good” (Colleague)

All FLOs agreed the role had a definite place in the future of hospital care, but it needed proper formalisation and induction for the ward staff and consultants. Training was recommended for FLOs without a clinical background to support them engaging with low tier clinical updates. Alternatively, the role could be separated into different roles, one that focused on medical updates and another which could focus on patient support.

> “If you’ve got a patient population that’s very vulnerable and very complex, and fairly elderly… those kind of wards is where they’ll be very valuable” (Colleague)

Clinical staff who work primarily in adult medicine were impressed with the system of care provided in paediatrics and how this could be transferrable to other areas of the hospital particularly elderly care, given the depth of research about how keeping patients stimulated (via music or art or other activities) can be beneficial (13).

> “I feel like it was like revolutionary geriatric care” (Colleague)

## Discussion

The current evaluation indicated that this was a role valued by healthcare professionals working in frontline COVID-19 areas where there were restrictions to visitors. Teams were unable provide the level of communication required by affected families and therefore the FLO role was implemented to ensure channels of communication were in place to prevent family distress and dissatisfaction. It was also to support clinical staff to reduce the moral distress of delivering patient care whilst not being able to provide a link to their families.

The role was implemented rapidly, with no policy in place so the nurses and youth worker/play specialists who acted as FLOs were initially unclear of their role; similarly, the healthcare team were also unsure of what the FLO did. However, over time as the FLO became embedded within the team, the benefit for acting as a conduit for information between the healthcare team and the family, and a link between patient and family became evident. An unexpected finding was the added benefits that those without a professional background brought to the role, providing stimulation to patients who would have otherwise been left to provide their own divertissement.

Effective communication is noted to be an essential element of family centred care and the family’s involvement in care is noted to be important for ensuring care meets the patient’s needs (14). This was recognised as a challenge during the pandemic as most healthcare organisations or government policies directed that visiting was greatly reduced or suspended. There are numerous papers that have described how organisations had implemented similar roles, providing details on the training for people in liaison roles and a checklist to guide how information was delivered (15-17). At a time when organisations were struggling to provide family support, this was invaluable but as we moved from the first to subsequent waves, it was important to determine whether this support adequately fulfilled the needs of families, patients and reduced burden to clinical teams. It was also important to provide clear definitions of the role to ensure those who were deployed into them were clear what was expected of them. However, to date, there has been limited evaluation of these roles and currently the focus has been on the clinical team’s perspective on how families were supported (18). The feeling of depersonalisation and moral distress nurses expressed at not being able to communicate effectively with families supports the implementation of these roles in times where visiting is restricted (18).

The current evaluation has a number of limitations. First, we were only able to engage five of the nine members of staff who had been in the FLO role. The four who were not represented may have not agreed to participate because they had a different perspective of the role that they were not willing to share so we could potentially have a biased view of the role.

Similarly, we were only able to interview seven colleagues who worked alongside the FLOs so there could be other views not captured in these data. Secondly, this evaluation focused on the FLOs who were based in COVID-19 ward areas not in critical care or high dependency units. These FLOs were mostly medical backgrounds, who have a different style of communication to nurses and youth workers (19). The types of information and manner in which they delivered this could therefore have been different. Finally, the benefit to patients and families is from the professional perspective which does not necessarily reflect the experiences of patients or families. This is something that warrants further investigation.

Despite these limitations, this evaluation indicated that there was benefit of providing a dedicated role to maintain and improve communication between healthcare teams and families during a crisis and is one of but a few emerging in response to COVID-19. While this is single centre evaluation, other organisations may recognise similar experiences so will be able to apply learning to their practices if the role is required in the future.

Based on the findings from the evaluation, we make the following suggestions for managing liaison roles if they are required in the future:

1. The FLO role should be divided into two tiers. One tier to provide clinical updates to families and liaising with the medical team. The other tier could focus on more pastoral care elements, more suited to care of the elderly, disabled adults or other vulnerable groups in long-term care. Restrictions on visiting are particularly difficult for these patients.
2. FLOs should be placed in an area in which they have had some experience. High dependency wards will need FLOs with more clinical knowledge in order to appropriately support families. These patients may not need as much engagement as the less sick patients, who need more pastoral attention to combat loneliness while visiting is restricted. Play specialists and youth workers seem best primed to providing pastoral care while those with a nursing, medical or allied health professional background are more appropriate for the higher need COVID wards.
3. The role needs more clearly defined boundaries and responsibilities. These boundaries need to be outlined through a proper induction and introduction to ward team, with a clear line of management on the ward. Medical staff should have the responsibility for communicating major or critical medical updates, while FLOs should provide updates to families on the day-to-day status/condition of the patient and help them to understand treatment plans.
4. Ethical issues related to the FLO’s boundaries need further discussion for their protection. FLOs should be available at the weekend and for longer periods of time during the day, similar to the shifts the clinical staff are working. This would require the ward to have a team of FLOs. However, it is not advised to rotate FLOs around different wards to cover shifts in order to preserve consistency of care.
5. Redeployment is a stressful period for everyone and all necessary steps should be taken to make this as smooth as possible to protect staff wellbeing (20).
6. Staff identified for redeployment as a FLO should also receive some training in communication skills, conflict resolution and resilience. FLOs should be give clinical supervision and brought together as a whole intermittently, this includes both tiers, to facilitate peer support and an opportunity to learn from one another.

The FLO role was introduced into clinical practice out of necessity and it is clear that the role should be a staple of pandemic protocols. However, there is compelling arguments for this to be a standard role within the NHS and not just during an emergency. Participants in the evaluation felt the role highlighted how regimented hospital care is when it comes to family communication and support. Carers tend to be left out of discussions and decision-making despite being an important element in patient’s treatment and recovery. Adapting this role outside of the pandemic could be an important step toward improved communication with patients and families and increasing the opportunity for them to be involved in decisions about their care.

## Data Availability

No data are available.

## Author contributions

All the authors were involved in developing the protocol. LH, LA co-ordinated the running of the study, LH was responsible for data acquisition. LH, RMT contributed to the analysis and drafted the manuscript. All authors critically revised and approved the final manuscript.

## Patient and public involvement

There was no patient and public involvement with this evaluation.

## Acknowledgements

We would like to thank our staff who took the time to participate in the study.

## Ethics approval

This project was assessed as being service evaluation according to the toolkit published by the English Health Research Authority (HRA). As such, no formal research ethics approval was required.

## Data sharing statement

No data are available.

## References

1. World Health Organisation. Coronavirus Disease (COVID-19) - events as they happen. Available from: https://www.who.int/emergencies/diseases/novel-coronavirus-2019/events-as-they-happen [accessed on 8th March 2021]

2. Hermann A, Deligiannidis K, Bergink V, Monk C, Fitelson E, Robakis T et al. Response to SARS-Covid-19-related visitor restrictions on labor and delivery wards in New York City. Archives of Women’s Mental Health. 2020;23(6):793–794.

3. Virani A, Puls H, Mitsos R, Longstaff H, Goldman R, Lantos J. Benefits and Risks of Visitor Restrictions for Hospitalized Children During the COVID Pandemic. Pediatrics. 2020;146(2):e2020000786.

4. Klompas M. Coronavirus Disease 2019 (COVID-19): Protecting Hospitals from the Invisible. Annals of Internal Medicine. 2020;172(9):619–620.

5. Boyle B. The critical role of family in patient experience. Patient Experience Journal. 2015;2(2):4–6.

6. Farrell M, Joseph D, Schwartz-Barcott D. Visiting Hours in the ICU: Finding the Balance among Patient, Visitor and Staff Needs. Nursing Forum. 2005;40(1):18–28.

7. Ottrey E, Palermo C, Huggins C, Porter J. Exploring staff perceptions and experiences of volunteers and visitors on the hospital ward at mealtimes using an ethnographic approach. Journal of Clinical Nursing. 2018;27(7-8):e1571–e1579.

8. Newell S, Jordan Z. The patient experience of patient-centered communication with nurses in the hospital setting: a qualitative systematic review protocol. JBI Database of Systematic Reviews and Implementation Reports. 2015;13(1):76–87.

9. Underwood J. Hospital visitors’ experiences at the nurses’ station. Nursing Standard. 2017;31(34):44–53.

10. Clay A, Parsh B. Patient-and Family-Centered Care: It’s Not Just for Pediatrics Anymore. AMA Journal of Ethics. 2016;18(1):40–44.

11. Health Research Authority. Is my study research? Available from: http://www.hra-decisiontools.org.uk/research/ [accessed on 8th March 2021].

12. Ritchie J, Spencer L. Qualitative data analysis for applied policy research” by Jane Ritchie and Liz Spencer in \edA.Bryman and R. G. Burgess [eds.] Analyzing qualitative data, 1994, pp.173–194.

13. Apóstolo J, Bobrowicz-Campos E, Gil I, Silva R, Costa P, Couto F, Cardoso D, Barata A, Almeida M. Cognitive Stimulation in Older Adults: An Innovative Good Practice Supporting Successful Aging and Self-Care. Transl Med UniSa. 2019; 6 (19):90–94. PMID: 31360672; PMCID: PMC6581488.

14. Heyland, D.K., Barwich, D., Pichora, D., et al., 2013. Failure to engage hospitalized elderly patients and their families in advance care planning. JAMA Intern. Med. 173 (9), 778–787. doi: 10.1001/jamainternmed.2013.180

15. Gabbi S, Man K, Morgan G,Maity S. The development of the family liaison team to improve communication between intensive care unit patients and their families during he COVID-19 pandemic. Arch Dis Child Educ Pract Ed, 2020, doi:10.1136/archdischild-2020-319726

16. Negro A, Mucci M, Beccaria P. Inroducing the Video call to facilitate the communication between health care providers ad families of patients in the intensive care unit during COVID-19 pandemia. Intensive and Critical Care Nursing, 2020, https://doi.org/10.1016/j.iccn.2020.102893

17. Lipworth AD, Collins EJ, Keitz SA et al. Development of a novel communication liaison program to support COVID-19 patients and their families. Journal of Pain and Symptom Management, 2021 61(1):e1–e10

18. Maaskant JM, Jongerden IP, Bik J et al. Strict isolation requires a different approach to the family of hospitalised patients with COVID-19: a rapid qualitative study. IJNS, 2021 https://doi.org/10.1016/j.ijnurstu.2020.103858

19. Adams A, Mannix T, Harrington A. Nurses’ communication with families in the intensive care unit - a literature review. Nurs Crit Care. 2017 Mar;22(2):70–80. doi: 10.1111/nicc.12141. Epub 2015 Jan 13. PMID: 25583405

20. San Juan, N. Camilleri, M, Jeans, J P, Monkhouse, A, Chisnall, G, & Vindrola-Padros, C. Redeployment and training of healthcare professionals to Intensive Care during COVID-19: a systemic review. MedRxiv. 2021;.01.21.21250230; doi: https://doi.org/10.1101/2021.01.21.21250230

